# TREATMENTS, RESOURCE UTILIZATION, AND OUTCOMES OF COVID-19 PATIENTS PRESENTING TO EMERGENCY DEPARTMENTS ACROSS PANDEMIC WAVES: AN OBSERVATIONAL STUDY BY THE CANADIAN COVID-19 EMERGENCY DEPARTMENT RAPID RESPONSE NETWORK (CCEDRRN)

**DOI:** 10.1101/2021.07.30.21261288

**Authors:** Corinne M. Hohl, Rhonda J. Rosychuk, Jeffrey P. Hau, Jake Hayward, Megan Landes, Justin W. Yan, Daniel K. Ting, Michelle Welsford, Patrick M Archambault, Eric Mercier, Kavish Chandra, Philip Davis, Samuel Vaillancourt, Murdoch Leeies, Serena Small, Laurie J. Morrison, on behalf of the Canadian COVID-19 Rapid Response Network (CCEDRRN) investigators for the Network of Canadian Researchers and for the Canadian Critical Care Trials Group

**Affiliations:** Department of Emergency Medicine, University of British Columbia, Vancouver, Canada; Centre for Clinical Epidemiology and Evaluation, Vancouver Coastal Health Research Institute, Vancouver, Canada; Department of Pediatrics, University of Alberta, Edmonton, Canada; Department of Emergency Medicine, University of Alberta, Edmonton, Canada; Division of Emergency Medicine, University of Toronto, Toronto, Canada; University Health Network, Toronto, Canada; Division of Emergency Medicine, London Health Sciences Centre, London, Canada; Schulich School of Medicine and Dentistry, Western University, London, Canada; Division of Emergency Medicine, McMaster University, Hamilton, Canada; Hamilton Health Sciences, Hamilton, Canada; Department of Family Medicine and Emergency Medicine, Université Laval, Québec, Canada; Centre de recherche du Centre intégré de santé et de services sociaux de Chaudière-Appalaches, Lévis, Canada; Centre de recherche, CHU de Québec, Université Laval, Québec, Canada; VITAM (Centre de recherche en santé durable), Québec, Canada; Department of Emergency Medicine, Dalhousie Medicine New Brunswick, Saint John, Canada; Department of Emergency Medicine, Saint John Regional Hospital, Saint John, Canada; Department of Emergency Medicine, University of Saskatchewan, Saskatoon, Canada; Department of Emergency Medicine, University of Manitoba, Winnipeg, Canada; Section of Critical Care Medicine, University of Manitoba, Winnipeg, Canada; Department of Emergency Medicine, St Michael’s Hospital, Unity Health Toronto, Toronto, Canada

**Author notes:** **Corresponding Author:** Corinne M. Hohl, MD MHSc. **Funding:** The Canadian Institutes of Health Research (447679), Ontario Ministry of Colleges and Universities (C-655-2129), Saskatchewan Health Research Foundation (5357), Genome BC (COV024) and Fondation du CHU de Québec (Octroi No. 4007) provided peer-reviewed funding. The BC Academic Health Science Network and BioTalent Canada provided non-peer reviewed funding. These organizations are not-for-profit, and had no role in study conduct, analysis, or manuscript preparation. **Declaration of interests:** The study authors have no conflicts of interest to declare.

**Keywords:** COVID-19, coronavirus disease 2019, SARS-COV-2, resource utilization, patient outcomes, pandemic waves

## Abstract

**Background:** Treatment strategies for coronavirus disease 2019 (COVID-19) evolved between pandemic waves. Our objective was to compare treatments, acute care resource utilization, and outcomes of COVID-19 patients presenting to Emergency Departments across two pandemic waves.

**Methods:** This observational study enrolled consecutive eligible COVID-19 patients presenting to 46 Emergency Departments participating in the Canadian COVID-19 Emergency Department Rapid Response Network (CCEDRRN) between March 1 and December 31, 2020. We collected data by retrospective chart review. Our primary outcome was in-hospital mortality. We used logistic regression modeling to assess the impact of pandemic wave on outcomes.

**Results:** We enrolled 9,967 patients in 8 provinces, 3,336 from the first and 6,631 from the second wave. Patients in the second wave were younger, fewer met criteria for severe COVID-19, and more were discharged from the Emergency Department. Adjusted for patient characteristics and disease severity, steroid use increased (odds ratio [OR] 8.0; 95% confidence interval [CI] 6.4 – 10.0), while the use of invasive mechanical ventilation decreased (OR 0.5; 95%CI 0.4 – 0.6) in the second wave. After adjusting for differences in patient characteristics and disease severity, the odds of hospitalization (OR 0.7; 95%CI 0.6 – 0.8) and critical care admission (OR 0.6; 95%CI 0.4 – 0.7) decreased, while mortality remained unchanged (OR 1.0; 95%CI 0.7-1.4).

**Interpretation:** In patients presenting to Canadian acute care facilities, rapid uptake of steroid therapy was evident. Mortality was stable despite lower critical care utilization in the second wave.

**Trial Registration:** Clinicaltrials.gov, NCT04702945

## INTRODUCTION

COVID-19 continues to place a strain on acute care hospitals around the world. Early reports from the first wave of the pandemic were critical in allowing clinicians to gain an understanding of a new disease entity,(1–6) but reflected convenience samples of patients with more severe disease and typical presentations due to limited testing capacity.(7) Most studies omitted Emergency Department (ED) utilization,(1–6) even though EDs are the first point of contact in the acute care system for many with COVID-19, where critical admission and discharge decisions have to be made.

Early in the pandemic many patients were treated with experimental therapies such as hydroxychloroquine, chloroquine, ritonavir/lopinavir, or ivermectin based on anecdotal evidence or inconclusive observational studies, some of which have been disproven.(8–10) While high-quality randomized controlled trials identified effective therapies and clear indications for their use,(11–13) others remain unsupported by high quality evidence.(14–16) Understanding changing treatments and resource utilization patterns is important to understanding the uptake of evidence-based therapies into clinical practice, and evaluating resource utilization and patient outcomes over time. These observations may guide jurisdictions with continued resource allocation challenges in future pandemic waves.

The Canadian COVID-19 Emergency Department Rapid Response Network (CCEDRRN, pronounced “*sedrin”*) is a national collaboration that harmonized data collection on consecutive COVID-19 cases in 50 EDs across 8 provinces (https://canadiancovid19registry.org/).(17) CCEDRRN’s goal is to generate real-world high-quality observational studies to evaluate and inform the pandemic response. The main objective of this study was to compare treatments, acute care resource utilization, and outcomes of COVID-19 patients presenting to EDs across the first two pandemic waves.

## METHODS

### Design and Setting

This pan-Canadian observational study enrolled consecutive eligible COVID-19 patients who presented to the EDs of 46 participating acute care hospitals between March 1 and December 31, 2020.(17) The research ethics boards of participating institutions reviewed and approved the study protocol with a waiver of informed consent for patient enrollment. Patient partners with lived experience from geographically distributed locations across the country were engaged from study inception to completion. All study sponsors were not-for profit organizations, and had no role in study design, data collection, analysis, interpretation, or writing of this manuscript. All authors had access to study data, and vouch for this manuscript.

### Study Population

Research assistants screened institutional or provincial medical microbiology testing lists for nucleic acid amplification tests (NAATs) for severe acute respiratory syndrome coronavirus 2 (SARS-CoV-2) and lists of presenting complaints or discharge diagnoses for consecutive eligible patients.(17) We excluded data from two sites that were unable to initiate data entry in 2020, and two sites that were unable to demonstrate ≥99% compliance with patient enrollment to ensure an unbiased sample.

We included all COVID-19 patients presenting to the EDs of participating sites, who were seen by an emergency physician, and whose medical record review was complete at the time of the data cut (Figure 1). We excluded patients tested in the context of an elective admission as part of a pre-admission checklist (e.g., planned hip revision), who were never seen by an emergency physician (e.g., seen directly by a consultant), and those who acquired COVID-19 in-hospital.

**Figure 1.**
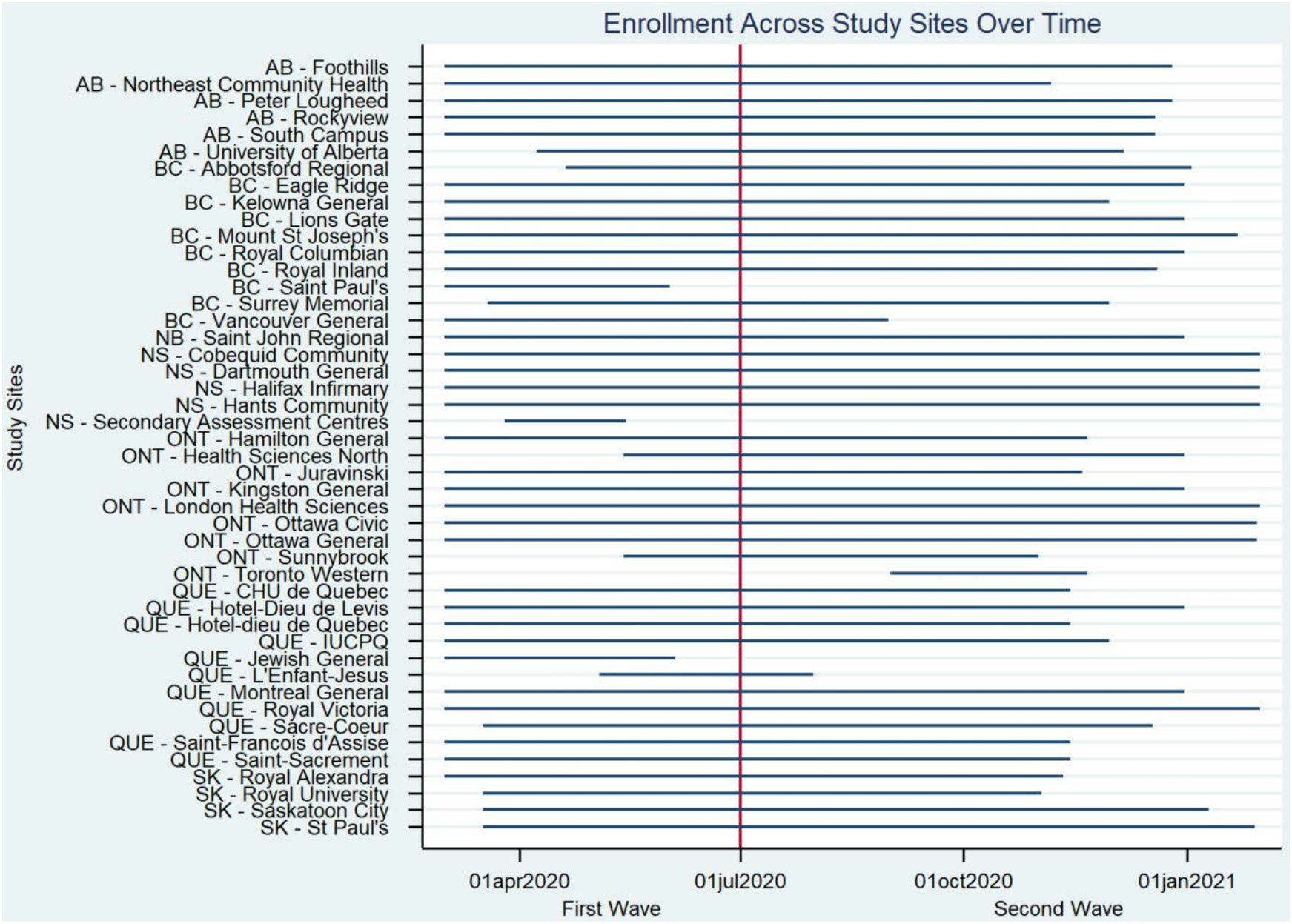
Gantt chart style for enrollment dates per site by pandemic wave. NS-Secondary Assessment Centre in NS closed in the first wave; ON-Toronto Western joined CCEDRRN in the second wave. We excluded four of 50 CCEDRRN sites. Two sites only started collecting data in 2021, and two sites had incomplete study trackers and were unable to demonstrate ≥99% compliance with patient enrollment.

### Definitions

We defined confirmed COVID-19 as patients presenting with ongoing COVID-19 symptoms and a positive nucleic acid amplification test (NAAT) for severe acute respiratory syndrome coronavirus-2 (SARS-CoV-2) obtained 14 days prior to, or after their ED arrival. This allowed us to capture patients who were diagnosed in the community and subsequently presented to the ED, and those with early false negative tests. We included patients presenting with COVID-19 symptoms and diagnosed with “confirmed COVID-19” to capture patients who were transferred into a CCEDRRN hospital whose NAAT at the sending site could not be confirmed, and patients who were presumed by treating clinicians to have COVID-19 despite negative NAATs.

We defined repeat COVID infections as cases in whom SARS-CoV-2 was isolated on two ED visits at least 90 days apart, based on reports of the longest duration of viral shedding reported.(18–20)

We defined a wave as a period of sustained acceleration in cases followed by a period of sustained deceleration in cases on the World Health Organization (WHO) dashboard for Canada. Based on this, we allocated patients to the first wave if they presented between March 1 and June 30, 2020, and to the second wave if they presented between July 1 and December 31, 2020.

We defined presentations for severe COVID-19 according to WHO age-based criteria.(21) For adults, criteria for severe COVID-19 were met if the patient had an oxygen saturation of <90% on room air, a respiratory rate >30 breaths per minute, or signs of severe respiratory distress documented in the ED medical record.

### Data Collection

Trained research assistants abstracted demographic and social, level of care, clinical, treatment, diagnostic and outcome variables from clinical records using standardized forms. We adhered to a data quality protocol and implemented data verification and quality checks to ensure high data quality.(17)

We calculated the seven-day moving average incident COVID-19 cases per 100,000 population for every health region included in the study.(22) We mapped every patient to the seven-day moving average incident COVID-19 case count of their health region using their postal code of residence and index ED visit date. As publicly available incident COVID-19 case data were not available for the early pandemic (0.1% of values were missing), we imputed values for the first five weeks of the pandemic by modeling reported COVID-19 over time using linear interpolation.(22)

### Outcomes

Our primary outcome was in-hospital mortality. Secondary outcomes included treatments, hospital and ICU admissions, and ED revisits and readmissions at seven and 30 days.

### Statistical Analysis

We summarized patient characteristics, treatments, and outcomes for each pandemic wave using descriptive statistics. We assessed wave differences with t-tests or analysis of variance (ANOVA) for continuous variables and chi-square tests for categorical variables. Separate logistic regressions with a random effect for patients modeled the associations between pandemic wave and the outcomes of interest. We considered different adjustments to provide an understanding of the incremental association between factors and the pandemic wave: (1) patient (age, sex, comorbidity, tobacco and illicit substance use) and presentation characteristics (arrival mode, arrival from, and WHO severe disease) recorded at the index ED visit, and (2) the variables in (1) as well as the seven-day moving average incident COVID-19 cases to account for changes in prognosis due to hospital burden.(23) We entered age and the seven-day moving average incident COVID-19 cases as continuous variables into our models; other variables were categorical. We conducted subgroup analyses on patients with severe COVID, pregnant patients, those reporting unstable housing, and those requiring invasive mechanical ventilation. We provided estimates with 95% confidence intervals (CIs). To ensure patient privacy, a cell size restriction policy prohibited us from reporting counts of less than five. A P-value less than 0.05 was considered statistically significant. We conducted all analyses using Stata (Version 16.1, StataCorp, College Station, Texas).

## RESULTS

### Main Results

We enrolled 9,967 COVID-19 patients, of whom 3,336 (33.5%) presented in the first and 6,631 (66.5%) in the second wave (Figures 1 & 2). Of these, 3,319 were enrolled in Quebec (33.3%), 2,868 in Alberta (28.8%) and 2,458 in British Columbia (25.6%). In all but 80 (0.8%) patients, a NAAT confirmed the COVID-19 diagnosis. Follow-up time was 30 days for discharged patients and between 30 and 229 days for admitted patients.

**Figure 2.**
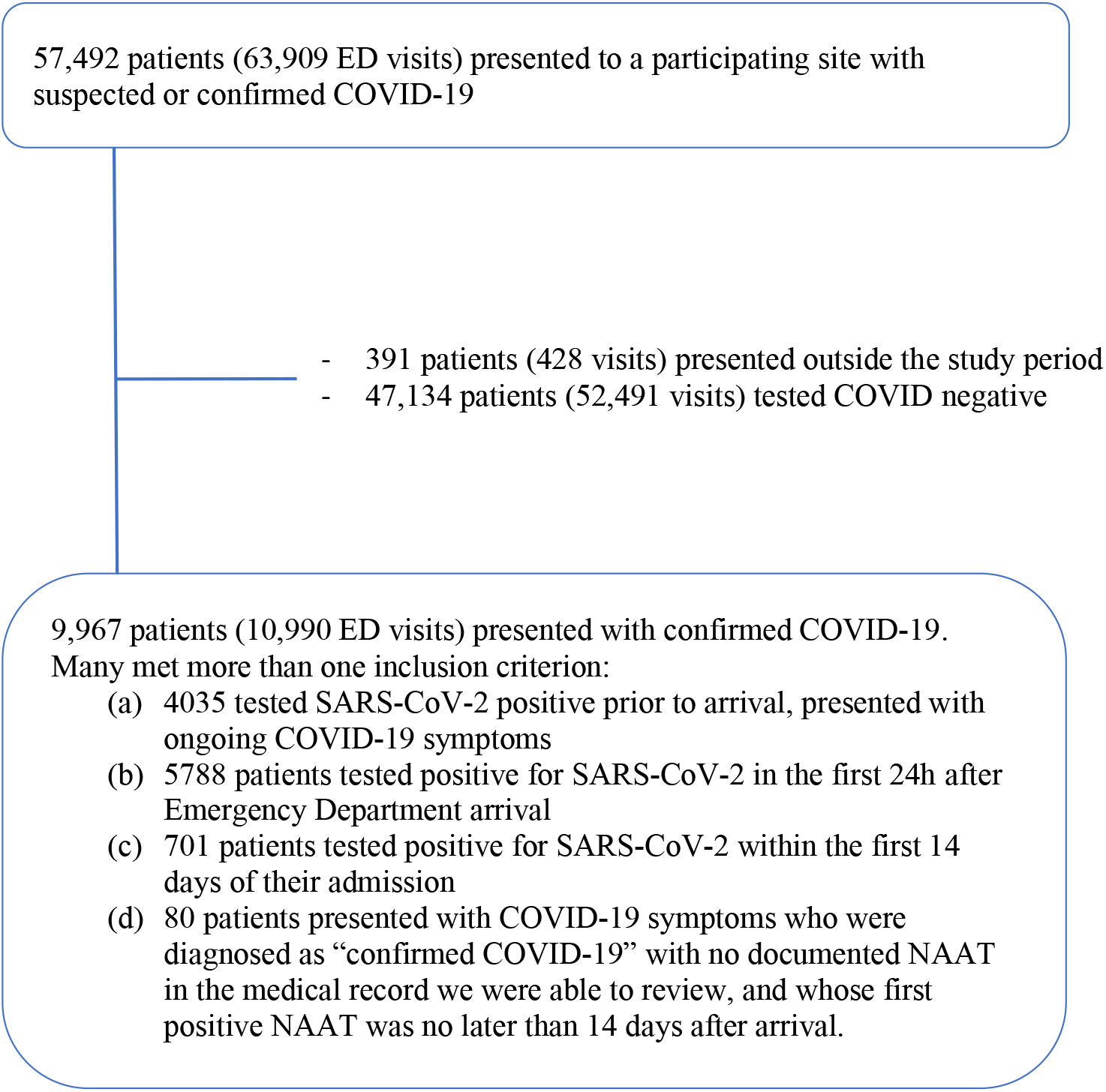
Patient flow diagram.

### Pandemic Waves

Patients presenting to acute care hospitals differed between waves (Table 1). During the second wave, patients were younger (mean age 53.2 versus 59.4 years old) with a similar proportion being female (49.2% versus 49.7%), and fewer comorbidities compared to the first wave. In the second wave, patients were less likely to arrive from long-term care (5.6% versus 18.3%), report an occupational exposure (2.3% versus 8.7%), travel-related infection (1.9% versus 6.8%) or an institutional exposure (7.5% versus 18.6%). Fewer patients met criteria for severe disease in the second wave (11.8% versus 17.0%).

**Table 1.** Patient and presentation characteristics by pandemic wave

| **Unique patients (=9,967)** | **First Wave**<br>**(n=3,336)** | **Second Wave**<br>**(n=6,631)** |
| --- | --- | --- |
| **Age (in years) mean (SD)** | 59.4 (20.7) | 53.2 (21.4) |
| **Age categories in years (%)** |  |  |
| < 1 | 6 (0.2) | 48 (0.7) |
| 1 – 9 | <5 | 52 (0.8) |
| 10 – 19 | 36 (1.1) | 157 (2.4) |
| 20 – 29 | 242 (7.3) | 780 (11.8) |
| 30 – 39 | 355 (10.6) | 919 (13.9) |
| 40 – 49 | 469 (14.1) | 1,008 (15.2) |
| 50 – 59 | 596 (17.9) | 1,070 (16.1) |
| 60 – 69 | 460 (13.8) | 898 (13.5) |
| 70 – 79 | 452 (13.6) | 821 (12.4) |
| 80 + | 716 (21.5) | 878 (13.2) |
| **Female (%)** | 1,657 (49.7) | 3,262 (49.2) |
| Pregnant (%) | 38 (1.1) | 79 (1.2) |
| **Arrival from (%)** |  |  |
| Home | 2,622 (78.6) | 5,941 (89.6) |
| Long-term care or rehab facility | 609 (18.3) | 373 (5.6) |
| Unstable housing* | 34 (1.0) | 136 (2.1) |
| Corrections | 6 (0.2) | <5 |
| Inter-facility transfer | 39 (1.2) | 66 (1.0) |
| **Goals of care (%)** |  |  |
| Full code | 2,584 (77.5) | 5,886 (88.8) |
| Intermediate goals of care | 344 (10.3) | 187 (2.8) |
| Do not resuscitate | 334 (10.0) | 526 (7.9) |
| **Risk for infection (%)** |  |  |
| Institutional (long-term care, corrections) | 662 (19.8) | 500 (7.5) |
| Unknown | 508 (15.2) | 2,158 (32.5) |
| Household contact | 421 (12.6) | 959 (14.5) |
| Occupational (healthcare worker) | 291 (8.7) | 154 (2.3) |
| Travel | 227 (6.8) | 126 (1.9) |
| **Comorbidities (%)** |  |  |
| Hypertension | 1,200 (36.0) | 1,830 (27.6) |
| Diabetes | 586 (17.6) | 1,051 (15.9) |
| Coronary artery disease | 292 (8.8) | 395 (6.0) |
| Asthma | 247 (7.4) | 463 (7.0) |
| Chronic lung disease, not asthma | 231 (6.9) | 343 (5.2) |
| Congestive heart failure | 129 (3.9) | 229 (3.5) |
| Active cancer | 121 (3.6) | 211 (3.2) |
| Obesity | 65 (2.0) | 126 (1.9) |
| Moderate / Severe liver disease | 15 (0.5) | 24 (0.4) |
| **Tobacco use (%)** | 92 (2.8) | 256 (3.9) |
| **Illicit substance use (%)** | 44 (1.3) | 181 (2.7) |
| **Unique ED visits (10,990)** | **(n=3,679)** | **(n=7,311)** |
| **Arrival by ambulance (%)** | 1,786 (48.6) | 2,963 (40.5) |
| **Canadian Triage Acuity Score (%)** |  |  |
| CTAS 1 (Resuscitation) | 186 (5.1) | 230 (3.2) |
| CTAS 2 (Emergent) | 1,039 (28.2) | 2,022 (27.7) |
| CTAS 3 (Urgent) | 1,876 (51.0) | 3,755 (51.4) |
| CTAS 4 (Less Urgent) | 498 (13.5) | 1,146 (15.7) |
| CTAS 5 (Non-Urgent) | 71 (1.9) | 150 (2.1) |
| **Arrival vital signs, mean (SD)** |  |  |
| Heart rate, beats per min | 93.7 (21.5) | 93.3 (19.2) |
| Systolic BP, mm Hg | 130.9 (21.6) | 130.8 (21.2) |
| Oxygen saturation, % | 95.3 (4.2) | 96.0 (3.7) |
| Respiratory rate, breaths per min | 21.7 (6.5) | 21.1 (6.5) |
| Temperature, degrees Celsius | 37.3 (0.9) | 37.0 (0.9) |
| **Symptoms reported at ED arrival (%)** |  |  |
| Cough | 2,152 (58.5) | 3,857 (52.8) |
| Dyspnea | 1,922 (52.2) | 3,626 (49.6) |
| Fever | 1,809 (49.1) | 2,822 (38.6) |
| General weakness | 1,049 (28.5) | 2,198 (30.0) |
| Chest pain | 887 (24.1) | 2,153 (29.4) |
| Diarrhea | 547 (14.9) | 1,002 (13.7) |
| Nausea/vomiting | 522 (14.2) | 1,359 (18.6) |
| Headache | 501 (13.6) | 1,266 (17.3) |
| Chills | 451 (12.3) | 1,289 (17.6) |
| Myalgia | 443 (12.0) | 1,163 (15.9) |
| Sore throat | 410 (11.1) | 900 (12.3) |
| Altered consciousness | 387 (10.5) | 561 (7.7) |
| Dysgeusia/anosmia | 132 (3.6) | 432 (5.9) |
| No symptoms | 125 (3.4) | 223 (3.0) |
| Pre-ED cardiac arrest | <5 | 9 (0.1) |
| **Symptom duration at time of the ED visit\*\*** |  |  |
| Mean (SD) | 6.0 (6.5) | 5.1 (5.3) |
| Median (IQR) | 4 (2 – 8) | 4 (1 – 7) |
| **WHO Severe Disease at ED arrival (%)\*\*\*** | 1,156 (31.7) | 2,026 (27.7) |

SD= standard deviation; CTAS=Canadian Triage Acuity Score; IQR=interquartile range;
ED=Emergency Department
\* Unstable housing includes homeless, shelter, single room occupancy
\*\* The denominator for symptom duration is 2,823 for wave 1, and 5,106 for wave 2.
\*\*\* We defined presentations for severe COVID-19 disease according to WHO age-based criteria. For adults, criteria for severe COVID-19 were met if the patient had an oxygen saturation of <90% on room air, a respiratory rate >30 breaths per minute, or signs of severe respiratory distress documented in the ED medical record.

Steroids were used more frequently (28.0% versus 9.5%, p<0.0001), and antimalarials (0.3% versus 9.0%, p<0.0001) and antivirals (1.5% versus 6.7%, p<0.0001) less frequently in the second wave (Table 2). Differences persisted after adjustment for baseline patient characteristics and disease severity (Tables 3a & b). A smaller proportion of patients were mechanically ventilated (3.7% versus 7.0%, p<0.0001) in the second wave versus the first, which also persisted after adjustment (OR 0.51; 95% CI 0.41 – 0.64). Even though patients were intubated at the same time after the onset of COVID-19 symptoms (6.5 versus 6.3 days, p=0.81, Appendix Table 1), they were intubated later in their hospital course (3.2 versus 2.0 days, p<0.0001) in the second versus the first wave, and for a shorter duration of time (12.8 versus 16.4 days, p=0.018).

**Table 2.** Acute care utilization and treatments of 9,967 patients, by pandemic wave

|  | **First Wave**<br>(n=3,336) | **Second Wave**<br>(n=6,631) | **P-value** |
| --- | --- | --- | --- |
| **Emergency department visits** |  |  |  |
| One ED visit (%) | 3,025 (90.7) | 6,039 (91.1) | 0.61<sup>t</sup> |
| Two ED visits (%) | 271 (8.1) | 526 (7.9) |  |
| Three or more ED visits (%) | 40 (1.2) | 66 (1.0) |  |
| **Admissions** |  |  |  |
| Never admitted (%) | 1,568 (47.0) | 4,078 (61.5) | <0.0001<sup>t</sup> |
| One admission (%) | 1,724 (51.7) | 2,481 (37.4) |  |
| Two admissions (%) | 40 (1.2) | 68 (1.0) |  |
| Three or more admissions (%) | <5 | <5 |  |
| Hospital days per admitted patients |  |  | <0.0001 |
| Mean (SD) | 15.6 (20.6) | 11.6 (12.0) |  |
| Median (IQR) | 8 (4 – 19) | 8 (4 – 15) |  |
| Admitted to critical care (%)* | 421 (12.6) | 510 (7.7) | <0.0001 |
| Critical care days per critical care admitted pts |  |  | <0.0001 |
| Mean (SD) | 15.6 (20.5) | 10.5 (11.3) |  |
| Median (IQR) | 10 (4 – 19) | 6 (3 – 13) |  |
| **Medication use (%)** |  |  |  |
| Steroids | 316 (9.5) | 1,854 (28.0) | <0.0001 |
| Antibiotics | 1,610 (48.3) | 2,368 (35.7) | <0.0001 |
| Antivirals | 219 (6.7) | 96 (1.5) | <0.0001 |
| Anticoagulation (heparin or oral) | 1,323 (39.7) | 2,119 (32.0) | <0.0001 |
| Antimalarials | 300 (9.0) | 21 (0.3) | <0.0001 |
| **Supplemental oxygen (%)** | 955 (28.6) | 1,124 (16.7) | <0.0001 |
| **Most aggressive form of oxygen delivery used\*\* (%)** |  |  |  |
| Mechanical ventilation (%) | 232 (7.0) | 247 (3.7) | <0.0001<sup>t</sup> |
| CPAP/BiPAP | 6 (0.2) | 18 (0.3) |  |
| High-flow nasal oxygen | 16 (0.5) | 52 (0.8) |  |
| Simple or non-rebreather facemask | 87 (2.6) | 103 (1.6) |  |
| Nasal prongs | 614 (18.4) | 704 (10.6) |  |

ED=Emergency Department; SD=standard deviation; CC=critical care; CPAP=Continuous Positive Airway Pressure; BiPAP=Bilevel Airway Pressure
\* Includes Critical Care, High Acuity/Step Down, and Operating Room (without surgery)
<sup>t</sup>ANOVA test for wave differences

**Table 3a.** Adjusted and unadjusted difference in therapy between 9,903 visits in wave 1 and wave 2^1^

| Treatments (%) | First Wave*<br>(n=2,690) | Second Wave<br>(n=7,213) | Unadjusted<br>Odds Ratio<br>(95% CI) | Adjusted<br>Odds Ratio§<br>(95% CI) |
| --- | --- | --- | --- | --- |
| Mechanical ventilation | 166<br>(6.2) | 245<br>(3.4) | 0.53<br>(0.44 – 0.65) | 0.53<br>(0.41 – 0.64) |
| Oxygen use | 620<br>(23.1) | 1,011<br>(14.0) | 0.54<br>(0.49 – 0.61) | 0.58<br>(0.48 – 0.71) |
| Steroid use | 201<br>(7.5) | 1,867<br>(25.9) | 4.76<br>(3.92 – 5.77) | 8.03<br>(6.41 – 10.04) |
| Antiviral use | 181<br>(6.7) | 96<br>(1.3) | 0.19<br>(0.15 – 0.24) | 0.15<br>(0.08 – 0.25)** |
| Anticoagulant use | 931<br>(34.6) | 2,133<br>(29.8) | 0.78<br>(0.71 – 0.87) | 1.00<br>(0.89 – 1.11) |
| Antimalarial use | 107<br>(4.0) | 22<br>(0.3) | 0.05<br>(0.02 – 0.11) | 0.03<br>(0.01 – 0.15) |

<sup>1</sup> We excluded 960 patients from 4 study sites that did not have enrollment in both waves.
§ Adjusted for age, sex, existing comorbidities (moderate or severe liver disease, hypertension, diabetes, congestive heart failure, coronary artery disease, asthma, chronic lung disease, active cancer, and obesity), WHO severe disease, arrival from, ambulance arrival mode, smoking status, and illicit substance use.
\* Reference category
\*\* Did not adjust for moderate or severe liver disease due to collinearity.

**Table 3b.** Adjusted and unadjusted difference in therapy between 2,986 visits with WHO severe disease on arrival in wave 1 and wave 2^1^

| Treatments (%) | First Wave*<br>(n=974) | Second Wave<br>(n=2,012) | Unadjusted<br>Odds Ratio<br>(95% CI) | Adjusted<br>Odds Ratio§<br>(95% CI) |
| --- | --- | --- | --- | --- |
| Mechanical ventilation | 125<br>(12.8) | 186<br>(9.2) | 0.69<br>(0.54 – 0.88) | 0.61<br>(0.47 – 0.78) |
| Oxygen use | 442<br>(45.4) | 690<br>(34.3) | 0.47<br>(0.35 – 0.63) | 0.52<br>(0.38 – 0.72) |
| Steroid use | 120<br>(13.2) | 1,061 (52.7) | 7.96<br>(5.29 – 12.00) | 9.51<br>(7.61 – 11.89) |
| Antiviral use | 94<br>(9.6) | 57<br>(2.8) | 0.27<br>(0.19 – 0.38) | 0.24<br>(0.17 – 0.34)** |
| Anticoagulant use | 495<br>(50.8) | 1,068 (53.1) | 1.09<br>(0.93 – 1.28) | 1.12<br>(0.95 – 1.33) |
| Antimalarial use | 56<br>(5.7) | 9<br>(0.5) | 0.07<br>(0.04 – 0.15) | 0.06<br>(0.03 – 0.12)** |

<sup>1</sup> We excluded 960 patients from 4 study sites that did not have enrollment in both waves.
§ Adjusted for age, sex, existing comorbidities (moderate or severe liver disease, hypertension, diabetes, congestive heart failure, coronary artery disease, asthma, chronic lung disease, active cancer, and obesity), arrival from, ambulance arrival mode, smoking status, and illicit substance use
\* Reference category
\*\* Did not adjust for moderate or severe liver disease due to collinearity

A greater proportion of patients were discharged directly from EDs in the second wave (61.3% versus 47.2%, p<0.0001; Table 4a). While a slightly higher proportion of patients revisited the ED within seven days (6.9% versus 5.8%, p=0.025), revisits were the same within 30 days (9.0% versus 8.8%, p=0.76) but more likely to result in admissions (8.2% versus 6.1%, p=0.008; Table 4b) in the second wave. In both waves a small proportion of patients died in the ED (0.5% versus 0.2%, p=0.016).

**Table 4a.**
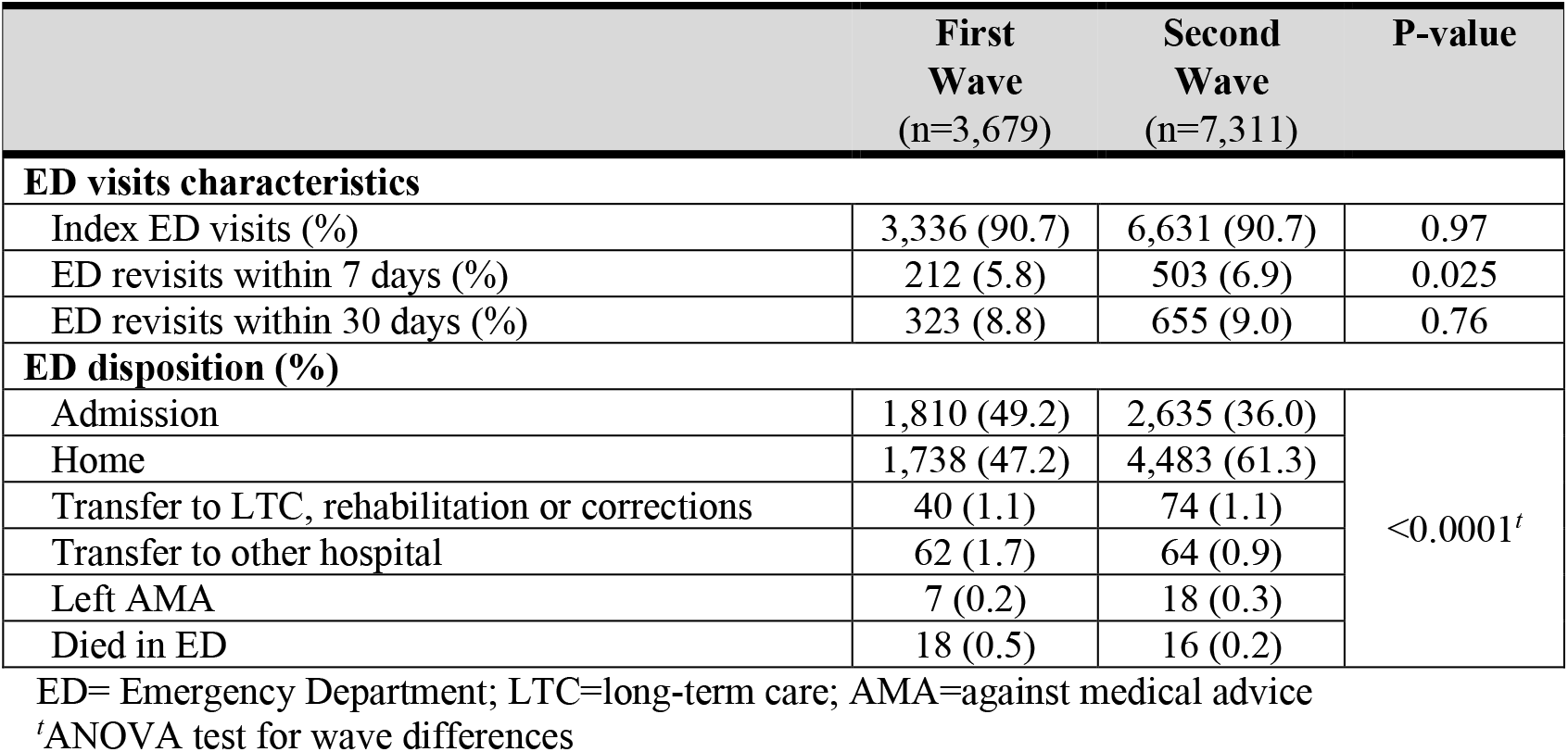
Emergency Department visits (n=10,990) by pandemic wave

**Table 4b.** Hospital admissions (n=4,445) by pandemic wave

|  | First Wave<br>(n=1,810) | Second Wave<br>(n=2,635) | P-value |
| --- | --- | --- | --- |
| Admission Characteristics (%) |  |  |  |
| Admission on index ED visit | 1,649 (91.1) | 2,330 (88.4) | 0.004 |
| Admission on ED re-visit within 7 days | 111 (6.1) | 217 (8.2) | 0.008 |
| Admission on ED re-visit within 30 days* | 153 (8.5) | 290 (11.0) | 0.005 |
| Level of Inpatient Care (%) |  |  |  |
| Ward only | 1,388<br>(76.7) | 2,123<br>(80.6) | 0.002 |
| Critical care* | 422<br>(23.3) | 512<br>(19.4) |  |
| Inpatient Trajectory (%) |  |  |  |
| From ED to ward | 1,388 (76.7) | 2,123 (80.6) | 0.001<sup>t</sup> |
| From ED to Critical Care** | 262 (14.5) | 283 (10.7) |  |
| From ED to ward to Critical Care** | 160 (8.8) | 229 (8.7) |  |
| Timing and Length of Admissions (%) |  |  |  |
| Admitted to ward on index visit | 1,243 (68.7) | 1,817 (69.0) | 0.004 |
| Admitted directly to Critical Care | 380 (21.0) | 456 (17.3) |  |
| Length of stay in hospital<br>Mean, (SD)<br>Median (IQR) | 15.6 (21.0)<br>9 (4 – 19) | 11.7 (12.0)<br>8 (4 – 15) | <0.0001 |
| Length of stay in Critical Care**<br>Mean, (SD)<br>Median (IQR) | 15.6 (20.5)<br>10 (4 – 19) | 10.5 (11.3)<br>6 (3 – 13) | <0.0001 |
| Died during hospitalization (%) | 346 (19.1) | 436 (16.6) | <0.0001 |

\*Includes 7-day readmissions
\*\*Includes high acuity/step down, and operating room for ventilation
<sup>†</sup>ANOVA test for wave differences

In the second wave, hospital admissions were shorter (mean 11.7 versus 15.6 days, p<0.0001; Table 4b), yet readmissions after hospital discharge were rare and similar across both waves (Appendix Table 2). In the second wave, fewer patients were admitted to critical care (7.7% versus 12.6%, p<0.0001; Table 2) and spent fewer days on average in critical care (10.5 versus 15.6 days, p<0.0001; Table 4b). These differences persisted after adjustment for differences in patient characteristics, disease severity, and the seven-day moving average incident COVID-19 cases (Table 5). Crude mortality was lower in the second wave (6.1% versus 8.5%; odds ratio [OR] 0.69, 95% CI 0.59-0.82), but disappeared after adjusting for patient characteristics, disease severity, and the seven-day moving average incident COVID-19 cases (OR 1.0; 95% CI 0.74-1.37).

**Table 5.**
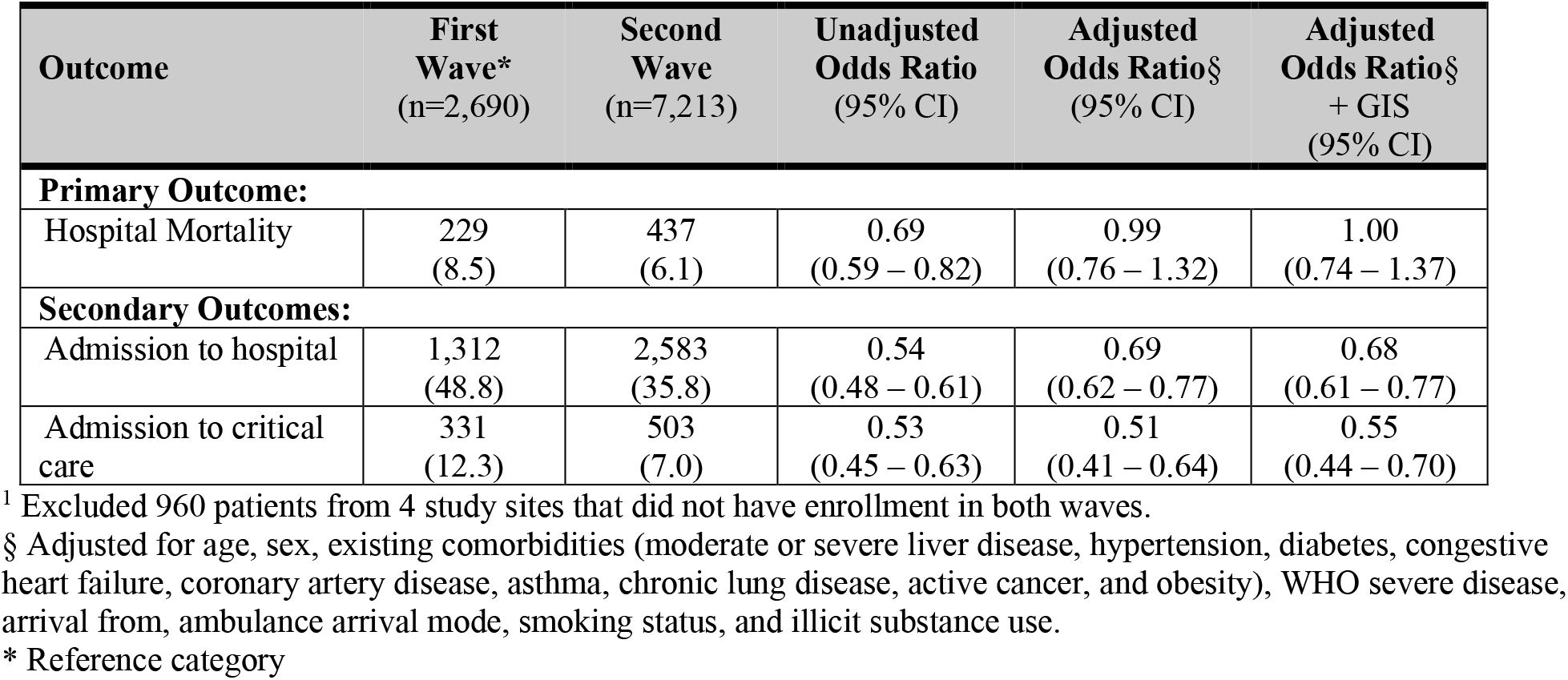
Crude and adjusted effect of pandemic period on the outcomes of 9,903 visits^1^

| **Outcome** | **First Wave\***<br>(n=2,690) | **Second Wave**<br>(n=7,213) | **Unadjusted Odds Ratio**<br>(95% CI) | **Adjusted Odds Ratio§**<br>(95% CI) | **Adjusted Odds Ratio§ + GIS**<br>(95% CI) |
| --- | --- | --- | --- | --- | --- |
| **Primary Outcome:** |  |  |  |  |  |
| Hospital Mortality | 229<br>(8.5) | 437<br>(6.1) | 0.69<br>(0.59 – 0.82) | 0.99<br>(0.76 – 1.32) | 1.00<br>(0.74 – 1.37) |
| **Secondary Outcomes:** |  |  |  |  |  |
| Admission to hospital | 1,312<br>(48.8) | 2,583<br>(35.8) | 0.54<br>(0.48 – 0.61) | 0.69<br>(0.62 – 0.77) | 0.68<br>(0.61 – 0.77) |
| Admission to critical care | 331<br>(12.3) | 503<br>(7.0) | 0.53<br>(0.45 – 0.63) | 0.51<br>(0.41 – 0.64) | 0.55<br>(0.44 – 0.70) |

<sup>1</sup> Excluded 960 patients from 4 study sites that did not have enrollment in both waves.
§ Adjusted for age, sex, existing comorbidities (moderate or severe liver disease, hypertension, diabetes, congestive heart failure, coronary artery disease, asthma, chronic lung disease, active cancer, and obesity), WHO severe disease, arrival from, ambulance arrival mode, smoking status, and illicit substance use.
\* Reference category

### Subgroups

During the study period, fewer than five of 9,967 patients (<0.05%, 95% CI 0.0002-0.0012%) re-visited a participating ED with a NAAT-confirmed re-infection greater than 90 days after their first visit. Among 119 pregnant patients, 28 (0.2%, 95% CI 0.2-0.3%) required admission, fewer than five (<0.05%, 95% CI: 0.013-0.09%) required mechanical ventilation, and none died. Among 176 (1.7%, 95% CI: 0.015-0.020%) patients reporting unstable housing (homeless, shelter, or single room occupancy), the proportion admitted was 50.6% (95% CI: 43.2-57.9%), and fewer than five (<5%, 95% CI: 0.84-5.93%) died.

## INTERPRETATION

### Main results

Our objective was to compare treatments, acute care utilization, and outcomes of COVID-19 patients presenting to acute care hospitals between pandemic waves. We found differences in patient characteristics between the first two waves reflecting public health measures to protect seniors and reduce travel.(24) We observed rapid uptake of evidence-based therapies and declining use of disproven therapies, indicating rapid translation of research evidence into practice. We observed decreasing hospital and critical care resource utilization over time, and less invasive mechanical ventilation with no adverse effect on mortality.

### Explanation of the findings

Unlike previous studies that were limited to single sites,(25–27) we enrolled patients in urban and rural, and academic and non-academic sites across Canada. We captured all COVID-19 patients, including vulnerable patients who are typically unable to provide informed consent (e.g., those with language barriers). Thus, we are confident that our sample is representative of COVID-19 patients who presented to Canadian EDs during the study period. We ascertained the outcomes of all enrolled patients, without censoring of patient outcomes at 28 or 30 days, or at the time of analysis, as was commonly done in early studies leading to incomplete outcome ascertainments.(14,28) We observed changes to the frequency, initiation, and duration of invasive mechanical ventilation over the study period associated with decreasing critical care resource utilization, consistent with other studies.(29) Early in the pandemic, non-evidence based criteria had been widely disseminated recommending early endotracheal intubation.(30) These recommendations were widely adopted despite lack of supporting evidence. While ventilation strategies continue to lack high-quality supportive evidence, this guidance has been questioned.(14–16) Our study does not allow for causal inferences, but documented less frequent invasive mechanical ventilation, later intubations, and a shorter duration of invasive mechanical ventilation in the second wave. These were associated with reduced critical care resource utilization and no adverse impacts on mortality.

In contrast to other studies, mortality was stable in our cohort after adjustment for differences in baseline patient characteristics and disease severity. Some studies that used administrative data observed decreasing mortality in Spring 2020, before any evidence-based treatments had been identified.(6,31) While some hypothesized that these observations were the result of improved clinical care as clinicians gained experience treating COVID-19, it is possible that these findings were the result of ascertainment bias.(7) Testing restrictions during the first wave meant that only the sickest COVID-19 patients were recognized and tested, which could have introduced systematic error.(32) Studies describing risk factors for mortality have consistently pointed toward age and respiratory parameters as the two most important predictors for deterioration and mortality in COVID-19.(28) Administrative database studies are unable to capture these clinical variables, and thus are unable to adjust for differences in disease severity at presentation. As a result, it is possible that ascertainment bias and residual confounding explain the early drop in mortality observed in administrative database studies.(6) In the early pandemic, residents of long-term care were tested more liberally than healthier populations. Oversampling of long-term care residents early on may have increased the observed mortality risk in the first wave compared to the second due to competing risks.(31) These differences may explain the observed differences in mortality trends across studies. In contrast, CCEDRRN sites were able to enroll consecutive patients across both waves when testing resources were adequate and used clinical data to adjust for baseline differences.

### Future directions

We were unable to link our data with genomic data to identify variants of concern which may be associated with higher in-hospital mortality.(33) While, variants of concern were limited in Canada during the study period, we plan to investigate this in future studies.

### Limitations of the study

We captured data retrospectively, and thus were limited to what was documented in medical records. We validated retrospectively collected data elements at several sites by comparing retrospectively collected data with prospectively collected data.(17) We were unable to link our data with genomic data to identify variants of concern which may be associated with higher in-hospital mortality.(33) However, there was limited circulation of variants of concern in Canada during the study period. We removed data from four sites making the study less generalizable, but instead ensured the integrity of our sample of consecutive cases to avoid selection bias.

## Conclusion

Our study documents rapid uptake of evidence during the COVID-19 pandemic, both for proven and disproven therapies, and efficiencies in resource utilization over time with increased rates of ED discharges and lower hospital and critical care resource use over time. This, indicates that advances in clinical decision-making and treatments created efficiencies, allowing health systems to safely care for greater numbers of patients.

## Data Availability

The CCEDRRN network endorses the guidance put forth by the World Health Organization to enable data sharing to optimize learning. CCEDRRN accepts applications for access to data by external investigators, prioritizing data requests by network Members.

## Acknowledgements

We gratefully acknowledge the assistance of Ms. Amber Cragg in the preparation of this manuscript. We thank the UBC clinical coordinating centre staff, the UBC legal, ethics, privacy and contract staff and the research staff at each of the participating institutions in the network outlined in the attached Supplement. The network would not exist today without the dedication of these professionals.

Thank you to all of our patient partners who shared their lived experiences and perspectives to ensure that the knowledge we co-create addresses the concerns of patients and the public. Creating the largest network of collaboration across Canadian Emergency Departments would not have been feasible without the tireless efforts of Emergency Department Chiefs, and research coordinators and research assistants at participating sites. Finally, our most humble and sincere gratitude to all of our colleagues in medicine, nursing, and the allied health professions who have been on the front lines of this pandemic from day one staffing our ambulances, Emergency Departments, ICUs and hospitals bravely facing the risks of COVID-19 to look after our fellow citizens and after one another. We dedicate this network to you.

**Appendix Table 1.**
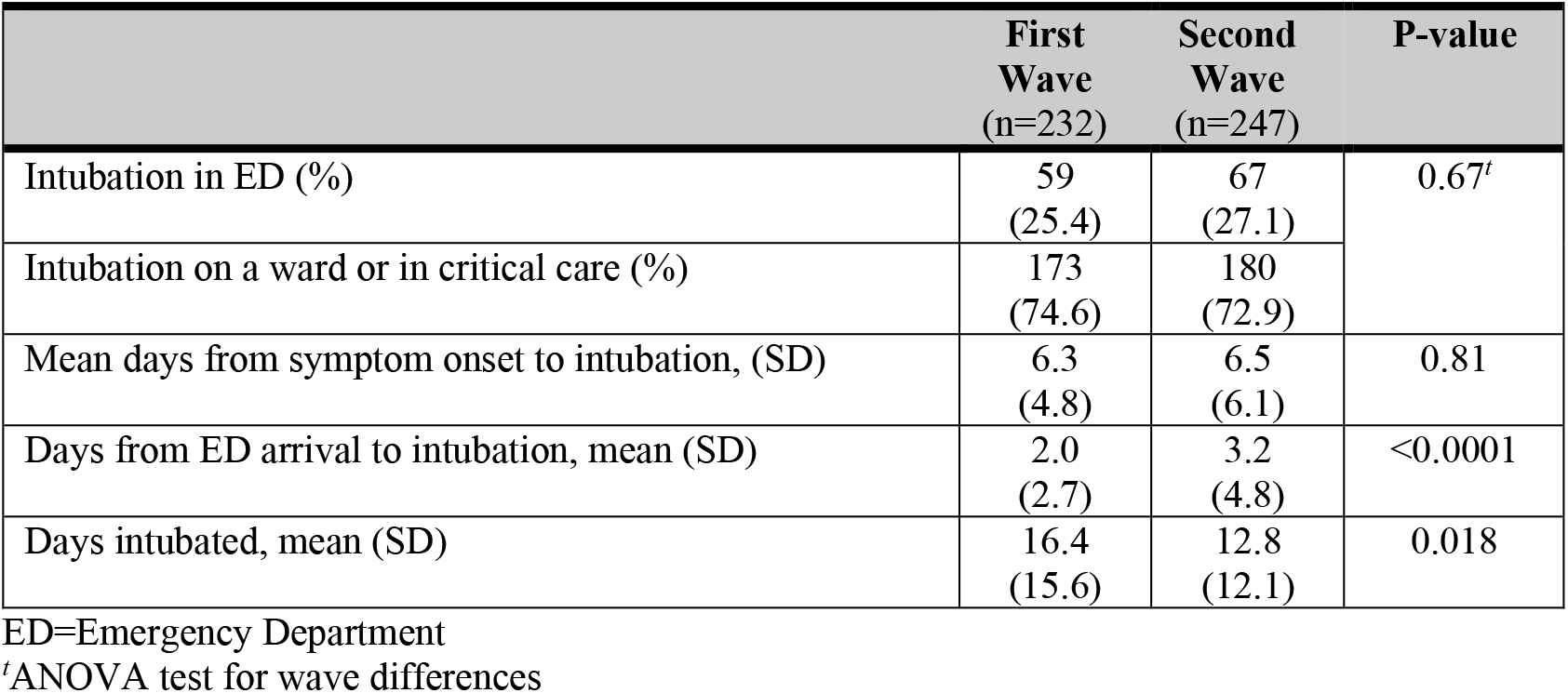
Characteristics of 479 mechanically ventilated patients

**Appendix Table 2A.** Number of hospital readmissions after index Emergency Department visits from which patients were discharged (n=361)

|  | **First Wave**<br>(n=124) | **Second Wave**<br>(n=237) | **P-value** |
| --- | --- | --- | --- |
| **Readmissions after index ED discharges (%)** |  |  |  |
| Readmission to ward within 7 days | 70<br>(56.5) | 152<br>(64.1) | 0.15 |
| Readmission to ward within 30 days* | 84<br>(67.7) | 186<br>(78.5) | 0.026 |
| Readmission to critical care within 7 days | 32<br>(25.8) | 35<br>(14.8) | 0.010 |
| Readmission to critical care within 30* days | 35<br>(28.2) | 43<br>(18.1) | 0.027 |

\*Includes 7-day readmissions

**Appendix Table 2B.** Number of hospital readmissions after index hospital admissions (n=106)

|  | **First Wave**<br>(n=38) | **Second Wave**<br>(n=68) | **P-value** |
| --- | --- | --- | --- |
| **Readmissions after discharge from index admission (%)** |  |  |  |
| Readmission to ward within 7 days | 6<br>(15.8) | 17<br>(25.0) | 0.27 |
| Readmission to ward within 30 days* | 26<br>(68.4) | 44<br>(64.7) | 0.69 |
| Readmission to critical care within 7 days | <5 | 8<br>(11.8) | 0.27 |
| Readmission to critical care within 30 days* | 7<br>(18.4) | 12<br>(17.7) | 0.92 |

\*Includes 7-day readmissions

